# A protocol to assess risk of fractures associated with use of menopausal hormone therapy: nested case-control study using CPRD

**DOI:** 10.1101/2024.05.15.24307422

**Authors:** Yana Vinogradova, Barbara Iyen, Tahir Masud, Lauren Taylor, Joe Kai

## Abstract

Menopausal hormone therapy (MHT) is prescribed to women with severe symptoms of menopause. Various studies have demonstrated increased bone density and decreased fracture risk in women using MHT. The randomised controlled trials were, however, run over only relatively short time periods, and there is no reliable or consistent evidence about fracture risk after MHT discontinuation. The proposed nested case-control study aims to fill this gap. We will use CPRD (GOLD and AURUM) and HES data. Every woman 40 years or over with a diagnosis of first fracture between 1998 and 2022 (a case) will be matched by age and general practice to up to 5 female controls with no previous records of fracture at the time of the case diagnosis (index date). MHT exposure will be assessed using prescriptions for the different types of MHT treatment historically available from the NHS. Conditional logistic regression will estimate fracture risk associated with duration of use and gap after discontinuation of MHT. The findings will be adjusted for smoking status, body mass index, family history of dementia, medical conditions and events, other medications and contraceptive drugs. A number of sensitivity analyses will be run to address the limitations of the study.

## Introduction

Osteoporosis and related fractures represent a significant burden in society, particularly for older women for whom risk of fracture increases with age, aggravated by menopause.^1^ Menopause is the stage of life when all women experience a drop in hormone levels in which oestrogen level is a key factor. Oestrogen deficiency affects the physiology of the whole body, and for some women this results not only in a range of unpleasant symptoms but also in a change in bone structure.

During menopause, bone mineral density (BMD) reduces each year by an average of about 1%, but for some groups the annual reduction can be between 2% and 5%.^2^ The risk of bone fractures, therefore, rises quickly in women after the age of 50. In women aged from 70 to 74, the risk of fractures is 8 times higher than for women aged from 50 to 54.^3^ Statistically, after the age of 50, every second woman will experience a fracture.^4^

Menopausal hormonal therapy (MHT) [also commonly known as hormone replacement therapy (HRT)] consists of an oestrogen (synthetic estradiol or conjugated equine oestrogen), normally in combination with a progestogen added to protect the womb. MHT is very effective in improving the quality of life for many women and has also been shown to maintain or improve bone health.

Older women, not on MHT and with an oestradiol serum level less than 5pg/mL, have been reported to have a risk of fractures 2.5 times higher than similar women whose BMD has been protected with oestrogen.^5^ A number of studies have also shown a reduced risk of osteoporosis among women currently taking MHT. Some of these studies have compared BMD levels in women using MHT and in controls who were not, and found in the control groups a rate of BMD decrease twice as high as that of women on MHT.^6 7^

Other studies have focused on fracture risks. Prior to the largest randomised controlled trial, the Woman’s Health Initiative (WHI), a meta-analysis had reported a 33% decreased risk of vertebral fractures and a 27% decreased risk of non-vertebral fractures in women randomised for MHT treatment.^8^ Although the study included a number of MHT formulations, only overall results were presented. The WHI trial itself was run on just two MHT treatments (conjugated equine oestrogen [CEE] alone and CEE in combination with medroxyprogesterone acetate [MPA]) and reported similar outcomes both for vertebral fractures ((HR 0.64, 0.44-0.93) and (0.68, 0.48-0.96) respectively) and for all fractures (0.72, 0.64-0.80 and 0.76, 0.69-0.83).^9^ The largest observational study to date, the Million Women’s Study, has also shown a decreased risk of fractures in women taking MHT (RR, 0.62; 95% CI, 0.58-0.66).^10^

Over the years many studies have confirmed the protective role of oestrogen in MHT treatments on the risk of fractures. A meta-analysis of 28 randomised controlled studies reported the following outcomes for current MHT users: decreased risk of all fractures (RR 0.74; 95% CI 0.69–0.80), decreased risk of hip fractures (RR 0.72; 95% CI 0.53–0.98); decreased risk of vertebral fractures (RR 0.63; 95% CI 0.44– 0.91).^11^ A recent meta-analysis of 12 observational studies on fragility hip fractures among MHT users was based on 305,879 cases and also demonstrated a decreased risk of hip fractures associated with MHT treatments (pooled odds ratio 0.80, 95%CI 0.65-0.98).^12^

However, although the risks of osteoporosis and of fractures have both been shown to go down, there has been significant inconsistency in findings. One study reported that BMD was declining at a similar rate both for treated and untreated women.^13^ By contrast, a Danish study, which assessed women who had completed 2-3 year courses of MHT, reported higher BMD for these patients than in their controls, not only at the end of their treatments but also in subsequent years.^14^ However, an American study, which followed 54,209 women for 6.5 years, reported that women who had discontinued were at 55% greater risk of a hip fracture than those still on MHT, with the risk increasing two years after cessation of treatment.^15^ The WHI trial reported an overall decreased risk of fractures after 13 years (hip fracture HR 0.91, 95% CI 0.72-1.15 (CEE); HR 0.81, 95% CI 0.68-0.97(CEE+MPA)).^9^ The National Osteoporosis Risk Assessment (NORA) study, which followed 140,584 women, found that hip fracture risk was similar between women who had discontinued MHT use for more than 5 years and women who had never used MHT, but that within 5 years of discontinuation the risk was *increased* in past users compared to non-users.^16^

Our recent population-based studies on risks of serious side-effects (VTE or breast cancer) in women using MHT have demonstrated that levels of risk differed between treatment regimens.^17 18^ Compliance to MHT treatments has been found to differ by progestogen type^19^ and we also found that risk levels can differ markedly between different MHT treatment formulations of oestrogen and progestogen. Currently available studies of fractures have either not distinguished between the outcomes for different MHT formulations or have been too underpowered to report such details. There is also little consistency both in risks found to be associated with different durations of MHT use, either before discontinuation or for current users,^20^ and in findings of how long the protective effect of MHT may stay after discontinuation.

In summary, current use of MHT is known to decrease the rate of fractures, but details about the differences in effect between different hormonal combinations in different treatments is still lacking. It is also known that the effect of MHT on fracture risk diminishes or disappears after discontinuation of treatment, but detailed evidence is contradictory. It is also not clear whether duration of reatment before discontinuation is important or whether patterns of decrease after discontinuation differ between different hormone combinations.

The many studies currently available differ widely in terms of their environment, data sources, size, selection criteria, coverage of MHT treatments and design. It would be useful, therefore, to have outcomes from a single environmentally- and design-coherent study, which includes the widest possible coverage of treatment formulations. This should be powered to deliver detailed levels of risk of fractures for MHT users in comparison with non-users, taking into account different treatment formulations and treatment regimens, both during use and after discontinuation.

The proposed study will use UK general population data routinely collected by general practices and hospitals, and will provide detailed estimates of fracture risks associated with different MHT use. It will include levels of risk of fracture for all available MHT treatments, by treatment type and duration, comparing MHT users with non-users. For each of the MHT treatments covered, the study will assess how the levels of risk for fractures change during treatment and after discontinuation. For different MHT combinations, the magnitudes of associations will be presented in terms of absolute numbers of patients. This detailed information will assist doctors and their patients to make more informed prescribing decisions and will also potentially contribute to the development of UK NICE guideline recommendations.

## Methods

### Source of data

The study will use routinely-collected data linked to hospital and mortality data from one of the largest UK primary care databases, CPRD. The study will use information collected using two computer systems, EMIS and VISION, which contribute data to the two subsections of CPRD, AURUM and GOLD. The collected information includes clinical values, diagnoses, referrals, laboratory and prescribing data, demographics and lifestyle measures. The database is representative of the UK general population, has been widely validated and is extensively used in epidemiological research.

We will use both GOLD and AURUM databases. Migrated practices will be used only once – the contribution to GOLD. The data in GOLD and AURUM were collected using different software (Vision and EMIS) so we plan to prepare the data separately on each part of CPRD using relevant sets of medical codes for extracting information. The resulting datasets will be identical both in coded variables and outcomes. There will be two underlying cohorts and two sets of case-controls.

### Study population

We will initially identify an open cohort of women aged 40 years and older registered with the study practices during the study observation period 1st January 1998 to 31st December 2022. The study entry date will be defined as at the latest of: the study start date (1st January 1998); practice up to standard date, date of registration with the practice plus 10 years, the woman’s 40th birthday. The cohort will be followed until the earliest of: the study end date (31st December 2022 or latest available); transfer out date; practice last collection date; patient death.

### Outcome

The outcome will be a record of fracture recorded in general practice, identified using the Read codes for fractures used in previous studies.^21^ Additional cases with incident fracture will be identified through linked hospital (HES) and mortality data using ICD10 codes of fracture recorded on hospital records or death certificates. Because not all fractures may occur as a direct consequence of osteoporosis but caused by a high impact accident, we will run a sensitivity analysis restricted to cases diagnosed with fragility fractures (hip, spine, rib, humerus, radius/ulna or pelvis).

### Selection of cases and controls

Cases will be women in the cohort who have a first ever record of any fracture in their records. We will match each case with up to 5 controls who are alive and registered with the same practice at the time of the fracture record of the case (index date) using incidence density sampling.^22^ Controls will be matched with cases by practice, age, and calendar time using incidence density sampling. Each control will be allocated an index date which will be the date of first diagnosis for the matched case. Cases and controls will only be included if they have at least 10 years of recorded data at the index date to ensure completeness of the records.

### Intervention

Information on all prescriptions for MHT before the index date in cases and controls will be extracted. A woman will be defined as a user if she has had at least one prescription containing systemic (oral, subcutaneous or transdermal) oestrogen indicated for menopausal treatment. The MHT to be included have been identified using the British National Formulary section 6.4.1.

We will categorise MHT by type of oestrogen (conjugated equine oestrogen or estradiol); type of progestogen (medroxyprogesterone, dydrogesterone, norethisterone or levonorgestrel/norgestrel); regimen of use (oestrogen only (or unopposed oestrogen) or oestrogen combined with progestogen); route of delivery (oral or transdermal/subcutaneous); dose for oestrogen; duration; recency of use.

Women will be defined as users of oral preparations if they took a tablet formulation of MHT, and users of transdermal/subcutaneous preparations if they used a patch, gel formulation or injection of oestrogen with or without a progestogen. To account for more than one route for a treatment (such as a tablet and a patch) we will have a separate variable for each. There is no evidence of fracture risk associated with other routes of administration (such as vaginal cream or pessary), but they will be included into analysis for consistency and to provide further information on risks associated with these routes. Another hormonal drug used for treatment of menopausal symptoms – tibolone – will also be included as separate exposure. We will also include raloxifene because it could be prescribed to menopausal women to maintain bone health.

For oestrogen, the dose will be categorised into low dose (<= 0.625mg for oral equine oestrogen or <=1mg for oral estradiol or <= 50 micrograms of transdermal estradiol) and high dose otherwise. We will consider as medium dose across all relevant prescriptions for a woman if she was exposed to both high and low dosages.

Duration of use will be assessed by calculating the number of days of exposure. If the gap between the end of one prescription and the start of next is 90 days or less, we consider exposure as continuous^23 24^ and combine the duration of the prescriptions. We will classify duration as short-term (up to 1 year), medium-term (1 to 4 years), long-term (5 to 9 years) and very long-term (10 or more years).

To investigate the effect of withdrawal on fracture risk we will assess recency of use by calculating the gap in days between the estimated date for last use of MHT and the index date. We will categorise recency as current (discontinued within 1 year before the index date), recent use (discontinued between 10 and 1 years before the index day) or past use (last use earlier than 10 years before the index date).

We will assess exposure at different times by combining duration and recency. We will analyse exact duration for different categories of recency and exact time after discontinuation for different categories of duration of use. We will also provide findings for categorised exposure/gap for comparison with other studies and estimations of absolute risks in the categories. Findings for sensitivity and subgroup analyses will be presented for categories of exposure/gap.

We will provide information for overall MHT use, overall oestrogen-only therapy, overall oestrogen-progestogen treatment and tibolone. We will also present the information for most-commonly prescribed combinations: unopposed conjugated equine estrogen (CEE); unopposed estradiol; oral and transdermal use of unopposed estradiol; oestrogen combined with medroxyprogesterone; oestrogen combined with levonorgestrel/norgestrel; estradiol combined with norethisterone; estradiol combined with dydrogesterone; tibolone; oral and transdermal use of unopposed estradiol, oral and transdermal use of estradiol with norethisterone/norgestrel.

No use before the index date will be the reference category for all exposures.

A woman will be considered as an oestrogen-only user if she had prescriptions for systemic oestrogen and no prescriptions for progestogen after the start of MHT. Any previous use of progestogen will be included as a confounder. If a woman had prescriptions for combined systemic MHT preparations or progestogen after MHT start she will be considered as an oestrogen-progestogen user. A small proportion of women may switch from combined to oestrogen-only therapy because of hysterectomy. In such events, we will still consider a woman as an oestrogen-progestogen user, calculating her exposure and gap since discontinuation for the therapy but include her exposure to oestrogen-only therapy to the analysis. For overall use of MHT, we will assess full exposure regardless of switching therapies.

### Covariates

Potential confounders will be variables which are either risk factors for fracture or indications for MHT use.^1 12 25 26^

#### Patient characteristics

- self-assigned ethnicity (using HES and GP data)
- body mass index (in kg/m^2^)
- Townsend deprivation score (patient level and if not available practice level)
- smoking status (non-smoker; ex-smoker; light [1-9 cigarette/day]; moderate [10-19]; heavy [≥20] smoker)
- alcohol consumption (no use; light [1-2 units/day]; moderate [3-6]; heavy [≥7] use)
- family history of osteoporosis (yes/no)
- oophorectomy/hysterectomy (yes/no)
- symptoms of menopause (yes/no)

#### Chronic conditions (yes/no)

- alcoholism
- cancers
- cardiovascular disease
- chronic kidney disease
- chronic liver disease
- chronic obstructive pulmonary disease
- dementia
- endocrine diseases (diabetes, hyperthyroidism, hyperparathyroidism)
- gastro-intestinal disorders (Crohn’s disease, ulcerative colitis, coeliac disease, chronic pancreatitis)
- HIV
- hypertension
- osteomalacia
- osteoporosis
- Paget’s disease
- rheumatoid arthritis

#### Use of other medications prior to the index date (yes/no)

- systemic corticosteroids
- proton-pump inhibitors
- antidepressants
- antipsychotics
- antiparkinsonians
- benzodiazepines
- sedatives
- H2-antagonists
- anxiolytic drugs
- thyroid hormone
- medications prescribed for osteoporosis treatment (bisphosphonates, vitamin D, raloxifene, denosumab, teriparatide, calcitriol)

### Data/statistical analysis

The main analyses will be run on all practices contributing to CPRD. To assess crude incidence rate in unexposed women, we will use the study population cohort but exclude women with prescriptions for MHT before entry and follow the rest until their first prescription of MHT. We will calculate crude incidence of any fracture and osteoporotic or fragility fracture in the initial and unexposed cohorts by dividing the number of incident fracture cases in the cohort by the number of person years.

We will use conditional logistic regression in the nested case-control study to estimate odds ratios with 95% confidence intervals for the MHT related variables. We will calculate unadjusted odds ratios and odds ratios adjusted for potential confounding variables listed above. We will include a potential confounder if addition of it would change the odds ratio for MHT exposure by at least 1% in either database sample.

To account for missing values for smoking status, alcohol consumption and body mass index, we will use multiple imputation to create five imputed datasets with multiple chained equations, applying Rubin’s rules to combine effect estimates and standard errors^27^. The imputation model will include all potentially important covariates, outcome status, years of records^28^. To test our assumption that data were missing at random, we will run a sensitivity analysis using only records with complete data. Missing values for ethnicity will be treated as the category ‘non recorded’.

All data preparation including multiple imputation and data analyses will be initially run on each data set. The results will be reviewed, combined using a meta-analysis technique and presented as combined estimates.^29^ Because the same type of data is collected in the same health care system and we use the same study design for both datasets, we will apply a fixed-effect model. A sensitivity analysis using a random effect model will also be run.

For continuous variables, such as body mass index and MHT exposures and gaps in years, we will use fractional polynomials to describe relationships between the variables and fracture risk.^30^ We will identify the components using unadjusted analysis, firstly on GOLD and AURUM samples separately, and then repeat unadjusted analysis on the combined sample to select the powers. The coefficients for the components will be obtained using adjusted analyses separately for GOLD and AURUM samples and then the final estimates will be assessed using the meta-analysis technique.

To address a possible confounding by indication bias – women or doctors may decide on MHT use because of already existing problems with bone health – we will run a subgroup analysis on women excluding those with diagnosis of osteoporosis, osteomalacia, diagnosis of cancer, Paget’s disease, alcoholism, HIV or anti-osteoporotic medications recorded before the index date.

We will repeat the analyses restricted to subgroups of cases diagnosed with fragility fractures. In this subgroup, it is possible that women started taking MHT because of already existing bone-related issues, so we will run a sensitivity analysis excluding information recorded in a year prior to the index date. This will address a possible protopathic bias caused by possible exposure due to symptoms of developing osteoporosis. For these subgroups, we will also present our findings separately for different types of fractures: hip, humerus and rib fractures.

To address consistency in capturing MHT exposure in women with advanced age, we will run subgroup analyses for women younger than 80 years and 80 years or older at the index date.

To address a possible information bias a sensitivity analysis will be run on a subgroup of practices linked to HES and ONS data. We will also use patient-level Townsend deprivation score as a confounder for these analyses and the observational period will be restricted to the earliest end of linked data.

Using combined incidence rate in the unexposed cohorts and combined odds ratios we will estimate the effects of exposures to different types of MHT and for different subgroups of women. We will provide the estimates for overall MHT use and, separately, for oestrogen-only and oestrogen-progestogen therapies.

A 1% level of statistical significance will be used to allow for multiple comparisons. Stata v 18 will be used for all the analyses.

## Patient or user group involvement

In conversations with perimenopausal and menopausal women, a number have expressed concerns about the negative side-effects of MHT and the need for comprehensive information about the risks (detrimental or beneficial) associated with different treatments. Perimenopausal women have also noted that they will “have to take MHT because it reduces the risk of osteoporosis”. So these women would appreciate more consistent information about the extent of such benefits for different treatments, and better information on how protective effects persist or diminish on cessation of MHT. We will discuss the findings in the light of women’s experience in terms of their health, and in particular bone fragility.

## Limitations of the study design, data sources, and analytic methods

MHT is widely considered as a form of osteoporosis prevention. The diagnosis of osteoporosis is not consistently recorded and also a proportion of the general population is underdiagnosed. An osteoporotic fracture could be considered as a proxy for osteoporosis as a condition.

Selecting the codes for osteoporotic or fragility fractures may miss a proportion of patients with fractures where medical codes are not specific. To deal with this limitation we define our main outcome as any fractures and plan to run an additional analysis for cases with fragility fractures. The limitations of the study will include the lack of formal adjudication of the type of fracture because some medical codes for fracture are not specific. There might be some false positives for cases and some false negatives for controls. The likelihood of misclassification is much higher for cases than for controls because of the relatively low incidence of fracture in the general population. The recording of fractures in CPRD has been validated by other studies and considered as very accurate.^31^

Another limitation is the potential misclassification of exposure to MHT. Women can easily access MHT through online prescribing services without seeing their own doctor but at a cost more than three times the prescription fee. We also do not know with certainty whether a woman has filled a prescription or whether/when she started taking a prescribed medication. We do not see, however, any reason why this should differ between cases and controls. These two potential misclassifications are likely to be small but might shift the odds ratios towards unity.

As for any observational study we acknowledge the potential for residual confounding as a further limitation.

## Data Availability

To guarantee the confidentiality of personal and health information, only the authors can have access to the CPRD data during the study in accordance with the relevant licence agreements. CPRD linked data were provided under a licence that does not permit sharing.

